# Maternal Body Mass Index and the Risk of Early-Onset Group B Streptococcus Disease in Newborns: A Systematic Review and Meta-Analysis

**DOI:** 10.1101/2025.08.10.25332710

**Authors:** Elisabeth Emanuel Graae, Niels Uldbjerg, Flemming Skjøth, Marie Aunskjær Bech, Pernille Nathalie Nielsen, Susani Rothmann Karkov, Caroline Margaret Moos, Mohammed Rohi Khalil

**Affiliations:** Department of Gynecology and Obstetrics, Lillebælt University Hospital, Kolding, Denmark; Department of Obstetrics and Gynecology, Aarhus University Hospital, Skejby, Denmark; Department of Clinical Medicine, Aarhus University, Aarhus, Denmark; Department of Regional Health Research, University of Southern Denmark, Odense, Denmark; Research Support Unit, Lillebælt University Hospital, Vejle, Denmark; Department of Clinical Research, Sønderjylland University Hospital, Aabenraa, Denmark

## Abstract

**Background:** Early-onset group B Streptococcus disease (EOGBS) remains a leading cause of neonatal morbidity and mortality. While intrapartum antibiotic prophylaxis based on clinical risk factors is widely implemented for prevention, emerging evidence suggests that maternal body mass index (BMI) may be an independent risk factor. This systematic review and meta-analysis evaluated the association between maternal BMI and the risk of EOGBS, as well as related proxy outcomes, including intrapartum vaginal GBS (Group B Streptococcus) and rectovaginal/urinary GBS colonization before term.

**Methods:** We systematically searched MEDLINE, Embase, and Cochrane CENTRAL on the 5^th^ of August 2024, for studies examining the relationship between pregestational BMI and EOGBS or its proxy outcomes. Eligible studies included observational and interventional designs, excluding case reports and conference abstracts. Risk of bias was assessed using the QUIPS tool. Random-effects meta-regression and sensitivity analyses were performed.

**Results:** We identified 16 eligible observational studies reporting data from a total of 2,621,817 women, encompassing 77,809 cases. For the risk of EOGBS and its proxy outcomes, our meta-regression showed a 2.7% increase in the odds ratio (OR) per unit increase in BMI. This corresponds to an OR of 1.4 (95% CI 1.1-1.7) for a BMI of 35 and 1.7 (95% CI 1.2-2.5) for a BMI of 45, compared to a normal BMI of 22. Sensitivity analyses by crude estimates showed an increase in the OR of 3.6% per unit increase in BMI, corresponding to an OR of 2.2 (95% CI 1.4-3.6) for a BMI of 45. One very large study on 1,971,346 live singleton births with 780 EOGBS cases, found a hazard ratio of 2.4 (95% CI 1.7-3.4) for a BMI of 35.0–39.9 compared to normal BMI.

**Conclusions:** Although the overall association appears modest, incorporating BMI may enhance prevention strategies for EOGBS.

This study protocol was registered in PROSPERO (CRD42023439201).

## 1. Background

Group B Streptococcus (GBS), transmitted vertically during labor, is a major pathogen responsible for potentially lethal early-onset neonatal infections (EOGBS), including sepsis, meningitis, and pneumonia [1]. EOGBS can be prevented by intrapartum antibiotic prophylaxis (IAP) administered to women colonized with vaginal Group B Streptococcus (vGBS) [1]. There are two main strategies for identifying candidates for IAP: a universal screening-based approach, where all pregnant women are screened for GBS colonization (typically at 35–37 weeks’ gestation), and a risk-based approach, which offers antibiotics only to women with predefined risk factors. The risk-based approach, used in several European countries including Denmark, relies on risk factors including a history of EOGBS in a previous infant, GBS bacteriuria during the current pregnancy, preterm labor, and prolonged rupture of membranes. However, EOGBS continues to occur among mothers who are not identified by these criteria, and up to 80% of EOGBS cases occur in neonates born to women without traditional risk factors [2,3]. Thus, ongoing discussions are taking place about how to enhance current screening and prevention strategies.

Although EOGBS is rare, with an incidence of 0.75 per 1000 livebirths in populations not offered IAP and 0.23 per 1000 livebirths in populations using IAP [4] GBS colonization in pregnancy is common, with a global prevalence of approximately 18% [5]. The predictive values of screening GBS cultures are critical to the time of assessment. Antepartum cultures are preferably obtained from both the vaginal introitus and anorectum, as emphasized in the AAP and CDC recommendations [1]

While established clinical risk factors for EOGBS focus on obstetric and infectious history, emerging evidence suggests that maternal metabolic factors may also influence colonization and transmission risk. In particular, increasing maternal body mass index (BMI) has gained attention as a potential risk modifier. Maternal obesity is known to impact immune responses, hormonal regulation, and microbiome composition, which may in turn affect susceptibility to infections. This includes not only urinary tract infections and wound infections, but also potentially asymptomatic colonization with pathogens such as GBS. Previous studies have reported an association between maternal BMI before conception and risks of EOGBS [6,7]. In addition, maternal BMI may be associated with EOGBS proxy outcomes, such as intrapartum vGBS and rectovaginal GBS colonization before term [7–11]. If these associations are substantial, high BMI should potentially be considered an additional risk factor for IAP eligibility. Therefore, this systematic review and meta-analysis aim to evaluate these associations.

## 2. Methods

### 2.1 Protocol and Registration

This study protocol was registered in PROSPERO (CRD42023439201) and conducted in accordance with the PRISMA 2020 guidelines.

### 2.2 Eligibility Criteria

We included original studies of any design (excluding case reports, conference abstracts and unpublished studies) that reported associations between maternal BMI assessed before 10 weeks of gestation and the outcomes of interest: EOGBS, intrapartum vGBS, or rectovaginal/urinary GBS colonization before term. There were no date or language restrictions.

### 2.3 Search Strategy

A comprehensive search was conducted in MEDLINE (Ovid), Embase (Ovid), and CENTRAL (Cochrane Library) on the 5^th^ of August 2024. Additional grey literature and citation tracking were performed using Google Scholar and Web of Science. Moreover, references cited by the included studies were also screened for eligibility. The grey literature and citation tracking did not yield any additional studies. The search strategy combined the terms: pregnancy, BMI/obesity, and GBS-related outcomes. Our search strategy included controlled vocabulary (MeSH) as well as free-text terms, combined in three blocks with the Boolean operator “AND.”. In each block, exploded medical subject headings or equivalents, depending on the database, truncations, free-text words, and narrower terms operators were used as appropriate. The following Medical Subject Heading terms were included in Embase and Medline; ***high risk pregnancy, pregnancy, pregnancy complications, childbirth, obesity, morbid obesity, body mass, body mass index, body mass, maternal obesity, streptococcal infections*** *and **streptococcus agalactiae**.* The search strategy is available in **S1 Appendix**.

### 2.4 Study Selection and Data Extraction

Two reviewers independently screened titles, abstracts, and full texts using Covidence. Discrepancies were resolved by consensus or third-party adjudication. Duplicates were removed using Covidence’s automated deduplication algorithm. Eligible studies were selected based on predefined PFO (Population, Factor, Outcome) criteria.

The PFO structure [12] was used to determine the suitability of article inclusion into the study and is available in the **S2 Appendix**. The primary outcome was EOGBS or its proxy outcomes, specifically rectovaginal or urinary GBS colonization.

- Population: Pregnant women.
- Factor: Pregestational BMI assessed before the 10^th^ gestational week.
- Outcome: EOGBS and its proxy outcomes, including intrapartum vGBS (vaginal Group B streptococcus), and rectovaginal or urinary GBS colonization before term.

Two reviewers independently extracted data in a predefined, partly adjusted form on study design, setting, BMI categorization, GBS detection methods, population demographics, and outcomes.

### 2.5 Statistical Analysis

The association between BMI and the risk of GBS across studies was analyzed using a random-effects meta-regression of the log odds ratio with standard errors estimated from the reported 95% confidence intervals. As the reported BMI intervals varied substantially between studies, we translated the intervals to a continuous scale by estimating the within-interval mean BMI through numerical integration, assuming a universal log-normal distribution with a mean of log(24.6) and a standard error of log(1.2) based on data from the Danish Medical Birth Registry. The reference categories representing normal weight were included with a low fixed standard error (0.001 on log scale). Reported adjusted odds ratios were used where available. Cohort studies reporting effects in terms of hazard ratios were included, assuming the hazard ratio approximates the odds ratio given the rare incidence of EOGBS. Due to overlapping cohorts in the two Swedish studies [6,7] evaluating BMI as an independent risk factor for EOGBS, the study with the shortest inclusion period, by Håkansson et al.[6] was excluded from the meta-analysis. Subgroup analyses on each of the proxy outcomes were performed, as well as pre-planned sensitivity analyses with restriction to results with OR<2, studies reporting adjusted OR, using only crude OR, and leave-one-out analyses. The estimated OR with 95% confidence intervals for the relative risk of outcome per one unit change in BMI was reported, as well as the association between BMI and OR of outcome was plotted with the average normal BMI as reference (BMI=22.3). Heterogeneity was assessed using Cochrane’s I² statistic. Publication bias was evaluated using funnel regression plots. *Stata Statistical Software: Release 18.5.* College Station, TX: StataCorp LLC was used for the analyses.

## 3. Results

### 3.1 Main results

844 studies were retrieved and imported into Covidence. The study selection process is documented in our PRISMA flow chart Fig 1. The most common reasons for exclusion were insufficient BMI data, e.g., only the mean BMI reported or a comparison of a BMI >23 (within the normal range) with a BMI <23, or a wrong study design, such as only an abstract being published. We included 16 eligible studies that reported data from a total of 2,621,817 women, encompassing 77,809 cases of either EOGBS or proxy outcomes (see Fig 1, Table 1). Of these, only two extensive Swedish register-based studies reported on EOGBS specifically[6,7]. Due to the overlap between the cohorts, only one of these studies [7] was included in the primary analysis. The remaining 14 studies reporting proxy EOGBS outcomes were categorized according to the timing of GBS assessment: intrapartum [13], gestational weeks 35 to 37 [11,14–16] (as performed in the universal routine screening in the US), or any other period during pregnancy [8–10,17–22]. Among the studies assessing GBS in gestational weeks 35 to 37, routine universal screening was established in 1 out of 4 study settings [11]. The studies included 8 European, 5 North American, one Asian, and two African populations, and all had either a retrospective or cross-sectional design. Study characteristics are presented in Table 1 and Table 2.

**Table 1.** Characteristics of included studies, geographical and temporal setup, size, design, and detection methods.

| Author, Year | Country | Study Period | N (Cases) | Study Design | Inclusion Criteria | GBS Detection Method |
| --- | --- | --- | --- | --- | --- | --- |
| Outcome: EOGBS |  |  |  |  |  |  |
| Villamor, 2021 (6) | Sweden | 1997-2016 | 1,971,346 (866) | Nationwide, multi-center cohort, | GA > 22 weeks, admission within 72 hrs | Blood culture-confirmed early-onset sepsis |
| Håkansson, 2008 (5)* | Sweden | 1997-2001 | 344,127 (248) | Nationwide, multi-center cohort, | GA ≥ 23 weeks, vaginal or emergency caesarean | Clinical diagnosis or blood culture |
| Outcome: Intrapartum GBS colonization |  |  |  |  |  |  |
| Khalil, 2022 (12) | Denmark | 2013-2014 | 902 (155) | Single-center, cohort | GA > 37 weeks, age >18, no antibiotics post-35 weeks | Rectovaginal culture PCR |
| Outcome: Antepartum GBS colonization week 35-37 (universal screening) |  |  |  |  |  |  |
| Alvareza, 2017 (10) | USA | 2013 | 2,045 (594) | Single-center, cohort | GA > 37 weeks, age 18-45, singleton pregnancy | Rectovaginal/ Urine culture |
| Rao, 2019 (13) | UK | 2014-2015 | 6,309 (1,836) | Single-center, cross-sectional | GA 35-37 weeks | Self-collected Rectovaginal culture |
| Rick, 2017 (14) | Guatemala | 2015 | 896 (155) | Single-center, cross-sectional | GA ≥ 35 weeks, age 15-45 | Rectovaginal culture |
| Namugongo, 2016 (15) | Uganda | 2015 | 309 (89) | Single-center, cross-sectional | GA ≥ 35 weeks, attended antenatal clinic | Anovaginal swabs, rapid GBS test kit |
| Outcome: Antepartum GBS colonization any time in pregnancy |  |  |  |  |  |  |
| Manzanares, 2012 (17) | Spain | 2007-2009 | 3,016 (399) | Single-center, cohort | Singleton pregnancy | Rectovaginal culture |
| Manzanares, 2019 (9) | Spain | 2012-2014 | 9,877 (1,835) | Single-center, cohort | GA > 26 weeks, singleton pregnancy, live births | Rectovaginal swab, GBS bacteriuria |
| Kwon, 2024 (21) | USA | 2010-2017 | 8,019 (997) | Single-center, cross-sectional | GA > 20 weeks | Rectovaginal culture |
| Dahan-Saal, 2011 (18) | France | 2004-2007 | 2,911 (cases) | Single-center, case-control | GA ≥ 24 weeks, singleton pregnancy | Vaginal/Urine culture, newborn sample |
| Kleweis, 2015 (16) | USA | 2004-2008 | 7,711 (1,989) | Single-center, cohort | GA > 37 weeks, singleton pregnancy | Rectovaginal culture |
| Venkatesh, 2020 (7) | USA | 2002-2008 | 115,070 (23,625) | Multi-center, cohort | GA ≥ 37 weeks | Urine, Vaginal, or Rectovaginal cultures |
| Stapleton, 2005 (8) | USA | 1997-2002 | 40,459 (cases) | Multi-center, case-control | Singleton pregnancy | GBS colonization from discharge record |
| Chen, 2018 (22) | China | 2015 | 2,121 (104) | Multi-center, cross-sectional | GA ≥ 28 weeks | Vaginal culture |
| Melchor, 2019 (19) | Spain | 2013–2017 | 16,609 (1,547) | Historical Single-center, cohort | GA ≥ 23 weeks, singleton pregnancy | Rectovaginal culture |

\*This study was not included in the primary analysis due to overlapping cohorts with Villamor et al, 2021 (6).
GA: Gestational age
GBS: Group B streptococcus
EOGBS: Early-onset group B streptococcus disease

**Table 2.** Characteristics of the study population, study population summaries

| Author, Year | Age (mean ± SD) | GA | GDM (%) | Adjusted covariates | Parity (P0 %) | Ethnicity | Socio-economic status | Smoking (%) |
| --- | --- | --- | --- | --- | --- | --- | --- | --- |
| Outcome: EOGBS. |  |  |  |  |  |  |  |  |
| Villamor, 2021 [7] | 30.1 (5.1) | 37-41: 88% | 1.6% | Maternal age, country of origin, education level, cohabitation with partner, parity, height, smoking, year of delivery | 58% | Nordic 80%, Non-nordic 19.9% | High: 37.9% $\geq 15$ years education | 8.7 |
| Håkansson, 2008 [6]* | NA | NA | NA | Maternal age, parity, GA, GDM | NA | Swedish population | NA | NA |
| Outcome: Intrapartum GBS colonization |  |  |  |  |  |  |  |  |
| Khalil, 2022 [13] | 87.1% age 25-40 | 37-40: 42.6%, $\geq 40$: 57.4% | NA | Parity, GA, vaginal delivery, smoking | 37.2% | Danish population | NA | 9.7 |
| Outcome: Antepartum GBS colonization week 35-37 (universal screening) |  |  |  |  |  |  |  |  |
| Alvareza, 2017 [11] | 27.9 ± 5.1 | 39.3 ± 1.3 | NA | Race, marital status, insurance, chronic hypertension, maternal genital infections | NA | White 26.5%, African American 48.7%, Hispanic 20.7%, Asian 2.6% | Marital status: Single 58%, Married 39.5% | 12.4 |
| Rao, 2019 [14] | 30.6 ± 5.6 | NA | NA | Race, parity, BMI, diabetes, GBS sample type | 46% | UK population | IMD Quintiles: 1-5 | NA |
| Rick, 2017 [15] | 25.4 ± 6.6 | $\geq 35$: 100% | NA | Maternal age, parity, education, marital status, literacy, ethnicity, home location, smoking, BMI, diabetes, prenatal care utilization, number of sexual partners, prematurity, previous infant with low birth weight or poor infant outcome | 32.5% | Non-indigenous 90.2%, Native Mayan 9.8% | Education: Primary 62.1%, Secondary 27.5% | NA |
| Namugongo, 2016 [16] | 25 | NA | NA | Maternal age, BMI, residence type, previous perineal tear | 37.2% | Banyankore 75.4%, Bakiga 9.7%, Baganda 12.3% | Education: Primary 34.3%, Secondary 38.8% | NA |
| BMI and risk of antepartum GBS colonization any time in pregnancy |  |  |  |  |  |  |  |  |
| Manzanares, 2012 [18] | 29.4 ± 5.2 | ≥37: 87.4%<br><33: 2.6% | 10% | Maternal age, parity, hypertension, diabetes | 43.1% | NA | NA | NA |
| Manzanares, 2019 [10] | 30.65 ± 5.68 | 276.7 ± 17.1 days | 5.9% | GA, parity, delivery mode, race and fetal birthweight | 34.7% | Caucasian 98.4%, Black 0.8%, Asian 0.6% | NA | 14.8 |
| Kwon, 2023 [22] | <20: 7.8%,<br>20-29: 52.1%,<br>30-40: 35.9%,<br>>40: 4.2% | ≥37: 77.5% | NA | BMI, maternal age, HSV diagnosis | 37.3% | Black 100% | NA | 3.3 |
| Dahan-Saal, 2011 [19] | 27.8 ± 6 | ≥37: 91% | 8.3% | Country of origin, prenatal care commitment | 39.8% | NA | Low: 69.3% ≤ high school, 45.2% unemployed | NA |
| Kleweis, 2015 [17] | 25.0 ± 6.1 | 39.0 ± 1.2 | NA | Race, parity, diabetes | 37.6% | Black 71.5%, Caucasian 19.6% | NA | 18 |
| Venkatesh, 2020 [8] | 27.2 ± 6.0 | 38.9 ± 1.1 | 4.2% | Maternal age, parity, pregestational diabetes, insurance status, study site, year of delivery | 6.3% | White 54.8%, Black 19.7%, Latina 19.3% | 63.2% had private insurance | 6.3 |
| Stapleton, 2005 [9] | 27.5 ± 6.2 | NA | 3.8% | Ethnicity, BMI, diabetes, parity, maternal age, alcohol, income, education, occupation smoking, prenatal care use | 44.4% | White 71.1%, Black 4.2%, Native American 2.5%, Hispanic 13.5% | High school: 52.2% | 11.5 |
| Chen, 2018 [23] | 27.7 ± 4.7 | 39.4 ± 1.4 | NA | BMI, GA, induced abortion, lotion use before pregnancy | NA | NA | Education: Junior high 38.5%, Senior high 31.9% | NA |
| Melchor, 2019 [20] | 33.9 ± 4.9 | 39.0 ± 1.9 | 5.3% | Maternal age, parity, hypertension, gestational age | 57% | NA | NA | 13.1 |

\*This study was not included in the primary analysis due to overlapping cohorts with Villamor et al, 2021 (6).
GA: Gestational age
GDM: Gestational diabetes mellitus
PE: Preeclampsia
NA: Not available
BMI: Body mass index
HSV: Herpes simplex virus
GBS: Group B streptococcus
EOGBS: Early-onset group B streptococcus disease

In the two Swedish nationwide cohort studies reporting EOGBS outcomes [6,7] the diagnosis was mainly based on culture-confirmed early-onset sepsis. The proxy outcomes used in the remaining studies were based on either intrapartum vaginal GBS-PCR [13], rectovaginal culture [14–18,20,22], vaginal culture [21] or mixed urinary/rectovaginal/vaginal GBS positive samples [8–11,19]. Common covariates included as potential confounders were maternal age, parity, smoking status, diabetes, race, socioeconomic status, hypertension, and preeclampsia (Table 2). The BMI categorization varied considerably across studies as illustrated in Table 3, which presents the studies’ main findings. When available, we based our analyses on adjusted outcome measures rather than crude measures, as emphasized in Table 3.

**Table 3.** Study-reported associations between BMI categories (intervals) and outcomes, with a summary of the study conclusions

| Study (Year) | Purpose | BMI Categories (%) | Main Results (Risk for GBS/EOGBS) | Summary Conclusion |
| --- | --- | --- | --- | --- |
| Outcome: EOGBS |  |  |  |  |
| Villamor, 2021 [7] | BMI & risk of early-onset sepsis (EOS) | <18.5: 2.2%<br>18.5-24.9: 64.6%<br>25-29.9: 22.6%<br>30-34.9: 7.5%<br>35-39.9: 2.3%<br>≥40: 0.8% | Adjusted HR for EOGBS:<br><18.5: aHR 0.73 (0.41-1.31)<br>25-29.9: aHR 1.2 (1.01-1.43)<br>30-34.9: aHR 1.7 (1.35-2.16)<br>35-39.9: aHR 2.4 (1.70-3.41)<br>≥40: aHR 2.56 (1.49–4.38) | Higher BMI increases risk of EOS. Strong dose-response, particularly for term infants. |
| Håkansson, 2008 [6]* | BMI & neonatal EOGBS | <18.5: 2.6%<br>18.5-24.9: 63.9%<br>25-29.9: 24.2%<br>≥ 30: 9.3% | aOR for culture-verified GBS septicaemia:<br><18.5: aOR 1.5 (0.6-3.8)<br>25-29.9: aOR 1.3 (0.9-2.0)<br>≥ 30: aOR 1.8 (1.1-3.0) | BMI ≥30 significantly increases risk of EOGBS. Effect is weaker but remains significant when including clinical diagnoses. |
| Outcome: Intrapartum GBS colonization |  |  |  |  |
| Khalil, 2022 [13] | BMI & intrapartum GBS colonization | < 18.5: 4.0%<br>18.5-24.9: 59.0%<br>25-29.9: 24.0%<br>≥ 30: 13.0% | < 18.5: aOR 0.26 (0.06-1.11)<br>25-29.9: aOR 1.62 (1.10-2.38)<br>≥ 30: aOR 2.68 (1.26-2.72) | Overweight and obesity are associated with increased GBS colonization, impacting the risk-based prevention strategy. |
| Outcome: Antepartum GBS colonization week 35-37 (universal screening) |  |  |  |  |
| Alvareza, 2017 [11] | BMI & GBS at 35–37 weeks | <25<br>25–29.9: 25.7%<br>30–34.9: 29.4%<br>35–39.9: 38.0%<br>≥40: 38.6 % | 25–29.9: aOR 0.97 (0.74–1.26)<br>30–34.9: aOR 1.14 (0.86–1.52)<br>35–39.9: aOR 1.43 (1.01–2.03)<br>≥40: aOR 1.27 (0.90–1.79) | Severe obesity appears associated with higher GBS risk, but the link weakens after adjusting for other variables. |
| Rao, 2019 [14] | BMI & GBS across racial groups | <18.5: 23.0%<br>18.5-24.9: 64.6%<br>25–29.9: 29.2%<br>30–34.9: 31.2%<br>≥35: 35.2% | <18.5: OR 0.76 (0.53-1.07)<br>25–29.9: OR 1.04 (0.92-1.18)<br>30-34.9: OR 1.15 (0.97-1.36)<br>≥35: OR 1.38 (1.09-1.74) | Highest GBS colonization rates found in Black women and those with higher BMI. |
| Rick, 2017 [15] | BMI & GBS across racial groups | <30: 69.6 %<br>≥30: 30.4% | ≥30: aOR 0.78 (0.52–1.18) | No significant association with BMI; GBS prevalence was high overall. |
| Namugongo, 2016 [16] | BMI & anogenital GBS | <25: 35.0%<br>25-29.9: 50.0%<br>≥30: 15.0% | 25-29.9: aOR 1.05 (0.58-1.9)<br>≥30: aOR 3.78 (1.78–8.35) | Obesity was the only significant predictor of anogenital GBS colonization. |
| Outcome: Antepartum GBS colonization any time in pregnancy |  |  |  |  |
| Manzanares, 2012 [18] | BMI & perinatal outcomes | <18.5: 5.5%<br>18.5-34.9: 86.2%<br>>35: 8.3% | <18.5: OR 0.72 (0.38-1.39)<br>>35: OR 1.57 (1.15–2.13) | Obesity increased GBS and perinatal risks; underweight linked to low birth weight and oligohydramnios. |
| Manzanares, 2019 [10] | BMI & GBS colonization | <25: 62.6%<br>25-29.9: 24.6%<br>>30: 12.8% | 25-29.9: aOR 1.05 (0.92-1.21)<br>>30: aOR 1.33 (1.12–1.56) | Obese women were 33% more likely to be colonized with GBS. EOGBS not directly evaluated. |
| Kwon, 2023 [22] | GBS & preeclampsia (Black women) | <25: 14.7%<br>25-30: 30.5%<br>>30: 54.8% | 25-30: OR 0.73 (0.6-0.89)<br>>30: OR 0.70 (0.58-0.84) | GBS-positive women had a significantly reduced risk of preeclampsia. |
| Dahan-Saal, 2011 [19] | BMI & vertical transmission | <18.5: 10%<br>18.5-24.9: 60.7%<br>25-29.9: 17.9%<br>≥30: 11.4% | Colonization:<br><18.5: aOR 0.81 (0.69-0.95)<br>25-29.9: aOR 1.06 (0.94-1.2)<br>≥ 30: aOR 1.19 (1.03-1.38)<br>Transmission:<br>OR 1.55 (1.16-2.06) | Obesity increases both GBS colonization and transmission risk to newborns. |
| Kleweis, 2015 [17] | BMI dose-response & GBS | <30: 41.8%<br>30-39.9: 43.4%<br>≥40: 14.8% | 30-39.9: aOR 1.29 (1.15-1.45)<br>≥40: aOR 1.54 (1.32–1.79) | Higher BMI showed a clear dose-response relationship with increased GBS colonization. |
| Venkatesh, 2020 [8] | BMI & GBS colonization | <25: 61.0%<br>25-29.9: 22.3%<br>30-34.9: 9.8%<br>35-39.9: 4.2%<br>≥40: 2.7% | 25-29.9: aOR 1.09 (1.05-1.13)<br>30-34.9: aOR 1.2 (1.15-1.26)<br>35-39.9: aOR 1.42 (1.33-1.51)<br>≥40: aOR 1.50 (1.38–1.62) | Risk of GBS colonization increases with each BMI category, especially in higher obesity classes. |
| Stapleton, 2005 [9] | Risk factors for GBS | <18.5: 4.3%<br>18.5-24.9: 54.6%<br>25-29.9: 23%<br>30-39.9: 15.2%<br>>40: 2.9% | <18.5: OR 0.98 (0.88-1.10)<br>25-29.9: OR 1.07 (1.01-1.12)<br>30-39.9: OR 1.20 (1.13-1.28)<br>>40: OR 1.45 (1.28–1.63) | Higher BMI is modestly but significantly associated with GBS colonization. |
| Chen, 2018 [23] | GBS in late pregnancy | ≤28: 98.7%<br>>28: 1.3% | >28: aOR 3.79 (1.28–11.26) | Strong association between BMI >28 and GBS in a Chinese cohort; small sample size. |
| Melchor, 2019 [20] | Obesity & perinatal outcomes | <30: 82.6%<br>≥30: 18.4% | ≥30: aOR 1.30 (1.14–1.47) | Maternal obesity increases risk of maternal and neonatal complications, including GBS. |

\*This study was not included in the primary analysis due to overlapping cohorts with Villamor et al, 2021 (6).
BMI: Body mass index
GBS: Group B streptococcus
EOGBS: Early-onset group B streptococcus disease

Our primary analysis presented in Fig 2 shows the OR of EOGBS or proxy outcomes as a function of BMI with normal BMI at 22.3 as reference with the shaded region representing 95% confidence interval based on random-effects meta-regression. Bubbles represent individual studies with diameters inversely proportional to the standard error of the reported association. Additionally, the individual results of two studies by Khalil et al.[13](intrapartum GBS colonization) and Villamor et al[7](EOGBS) are depicted. The meta-analysis showed an exponential increase in OR equal to an OR of 1.027 per BMI unit (Fig 2). When compared to a reference BMI of 22.3, this corresponds to an OR of 1.21 (95% CI 1.07–1.36) at BMI 30, 1.37 (95% CI 1.12–1.67) at BMI 35, 1.55 (95% CI 1.18–2.04) at BMI 40, and 1.75 (95% CI 1.23–2.49) at BMI 45.

In Fig 3, a forest plot is visualized, representing the estimated association between BMI and risk of EOGBS or proxy outcomes by odds ratios with 95% confidence intervals based on random-effects meta-regression for the primary analysis and various subsets of included studies. A sensitivity analysis was conducted on crude OR (Fig 3), showing an association between change in GBS rate and change in BMI (“Δ rate of GBS per Δ BMI”) of 1.036 (95% CI: 1.026–1.045), compared to the aOR of 1.027 (95% CI: 1.011–1.043). An OR of 1.036 per BMI unit corresponds to an OR of 1.31 (95% CI 1.12-1.54) at BMI 30, 1.57 (95% CI 1.21-2.03) at BMI 35, 1.87 (95% CI 1.30-2.69) at BMI 40, and 2.23 (95% CI 1.40-3.56) at BMI 45. Supplemental subgroup analyses on the studies assessing GBS at gestational weeks 35 to 37 or anytime in pregnancy weakened the association (Fig 3). However, the analysis on gestional week 35 to 37 studies showed a markedly lower I^2^ value of 17.5%, indicating less heterogenicity between these studies [11,14–16]

As indicated in Fig 2, two studies—both included in the primary analysis—demonstrated markedly stronger associations. One of these [7], assessed as having a low risk of bias, was based on a large Swedish cohort, and used EOGBS as the outcome. It reported an adjusted hazard ratio of 2.4 (95% CI 1.7–3.4) for BMI 35–39.9, with an even stronger association observed among term infants. The other study[13], assessed as having a moderate risk of bias, used intrapartum GBS colonization as outcome and reported an OR of 2.68 (95% CI 1.26-2.72) for a BMI ≥ 30. Despite this, the leave-one-out analyses did not reveal any crucial importance in the main analysis of any included study (**S4 Appendix**).

For the remaining proxy outcomes—based on four studies assessing GBS colonization at or after the 35^th^ gestational week [11,14–16] and nine studies assessing GBS status at any time during pregnancy [8–10,17–22] – only studies with limited weight in the primary analysis showed results that deviated significantly from the overall findings. This resulted in an I^2^ of 64% (Fig 2), which, however, may represent substantial heterogenicity among included studies.

### 3.2 Risk of Bias Assessment

The QUIPS (Quality in Prognosis Studies) tool was used to assess bias across six domains: study participation, attrition, prognostic factor measurement, outcome measurement, confounding, and statistical analysis. The risk of bias was evaluated and is available in Fig 4. The assessment was performed by two authors working independently.

According to the QUIPS assessment, most studies were rated as having a low risk of bias whilst some were rated as moderate, primarily due to issues related to study attrition, selection bias, or missing information on exposure and outcome characteristics (Fig 4). In general, detailed information on BMI assessment methods was limited, and pregestational BMI was often likely self-reported, introducing a risk of recall bias. A detailed description of GBS assessment, including the sample method, anatomic location, and distribution in cases with multiple locations, as well as microbiological analysis, was lacking in many studies. In Fig 5, a funnel plot is depicted for all included studies for assessment of publication bias. The dashed horizontal line shows the pooled OR and circles represent individual study estimates (log odds ratio) against the respective standard error. In conclusion, there was consistency in the distribution of the studies, with a few outliers, indicating a low risk of publication bias.

## 4. Discussion

### 4.1 Principal Findings

This systematic review suggests a dose-response relationship between increasing preconception BMI and increasing risk of a combined outcome, including EOGBS, intrapartum vGBS and rectovaginal GBS colonization. The dose-response relationship demonstrated by several studies, where the risk of GBS outcomes increased with each successive BMI category, was observed in all studies including more than one obesity class (defined by a BMI >30) [7–9,11,14,17].

Although the overall ratio was below 2, it is notable that it exceeded 2 for the most important outcomes: EOGBS and intrapartum GBS colonization, and that the results were consistent.

Regarding EOGBS, two extensive overlapping cohort studies—Villamor et al. and Håkansson et al.— demonstrated that women with higher BMI had significantly greater risk of delivering infants with EOGBS [8, 10]. Villamor et al. observed an adjusted hazard ratio of 2.56 for BMI ≥40, indicating a more than twofold increase in risk compared to women with normal weight. Håkansson et al., with a shorter study period, similarly found a significantly elevated adjusted odds ratio of 1.8 for women with BMI ≥30. Regarding intrapartum GBS, Khalil et al. found that overweight and obesity significantly increased the likelihood of intrapartum GBS detection, with adjusted OR of 1.62 (1.10-2.38) in overweight, and 2.68 (1.26-2.72) in obese women.

However, not all findings were uniform. Kwon et al. reported an inverse association [22] and Rick et al. and Rao et al. reported either non-significant or attenuated associations [14,15]. The BMI–GBS relationship may be influenced by contextual factors such as ethnicity, health system differences, or socio-economic status. For instance, Rao et al. found the highest colonization rates in Black women and those with higher BMI, raising the possibility of effect modification by race [14]. The association between Black people and GBS colonization and disease is consistent with other studies [24,25], moreover reporting an independent association with Black people [24,26]. However, these observational data do not allow elucidation of causality.

In this meta-analysis, we used the adjusted odds ratio (aOR) from publications that reported both crude OR and aOR. However, one could argue that many of the variables included in the adjustments (Table 2) are not true confounders but rather intermediaries or effect modifiers. To clarify this, a sensitivity analysis was conducted on crude OR showing a 1.036 (95% CI: 1.026–1.045) change in OR per BMI unit compared to the aOR of 1.027 (95% CI: 1.011–1.043) (Fig 4). Excluding potential intermediates or effect modifiers as when using the aOR may result in an underestimation of the BMI-associated risk. Therefore, using the crude OR might be more appropriate in a clinical setting.

The biological mechanisms linking increased BMI and GBS colonization are not yet fully understood, but several hypotheses exist. Obesity alters immune responses, gut and vaginal microbiota, and inflammatory pathways, all of which may predispose individuals to persistent GBS colonization [27]. Additionally, metabolic dysregulation common in obesity may impair mucosal defences, thereby facilitating GBS persistence and vertical transmission during labor. The association between obesity and EOGBS may involve specific pathways, including the interaction of dietary fats, such as palmitate (22) which can induce inflammatory responses in the gestational membranes and the placenta [28] potentially increasing susceptibility to GBS colonization [29].

Although BMI is not a standard risk factor in many existing clinical guidelines for EOGBS, both research and clinical practice support that high maternal BMI may increase the risk of EOGBS [30–32]. Besides obesity, GDM and pregestational diabetes are novel risk factors that have been studied. A recent meta-analysis from 2024 found GDM was associated with a 16% elevated risk of maternal GBS colonization, and an even larger risk of 76% reported for pregestational diabetes, resulting in a pooled OR of 1.76, CI 1.27–2.45 [33]. In the literature, increasing risk following tobacco use, Black individual, younger maternal age, and hypertension are other suggested risk factors for maternal GBS colonization [24,26]. There must be a correlation between some of these factors, as obesity, for example, correlates to an increased risk of diabetes and hypertension.

### 4.2 Clinical Implications

In settings where risk-based GBS screening is practised, maternal BMI could be considered as an additional risk factor. The strongest predictive value for EOGBS is positive GBS culture at delivery (OR 204.0) compared to a lower risk at 28 (OR 9.64) and 36 weeks of gestation (OR 26.7), outlining that intrapartum GBS assessment is ideal in a risk-based screening, especially with improved PCR technology allowing quick and accurate testing [1] In comparison, the traditional risk factors in the risk-based screening model are notably stronger than the BMI related risk found in this review; e.g. prolonged rupture of membranes > 18 hours (OR: 7.28), intrapartum fever or PROM at term (OR 11.5) and gestation <37 weeks (OR: 4.83) [1]. However, including BMI in risk stratification may reduce the number of missed cases, as well as decrease the use of unnecessary IAP. Therefore, it may be wise to combine a risk-based strategy with intrapartum GBS-PCR [34–36].

An OR of 1.036 means that for every unit increase in BMI, the risk of EOGBS or intrapartum GBS colonization increases by 3.6% (Fig 3). Although this increase may seem small, it is statistically significant and could have an impact, especially at a population level. If many pregnancies involve women with higher BMI, the overall risk of EOGBS could rise, even with a modest increase in BMI. In Denmark, for instance, women with a BMI > 35 account for 5.7% of the birthing population [37]. Moreover, women with a high BMI may face a 10% increased risk of a false-negative risk stratification [38,39].

Before revising the EOGBS prevention program, it is essential to consider all aspects of a medical technology evaluation. In addition to clinical effectiveness, these include safety (e.g., allergic reactions and microbiome alterations), risks (e.g., bacterial resistance to antibiotics), costs (both short- and long-term), implementability (e.g., training requirements for healthcare staff), patient perspectives, ethical implications (e.g., equity), and alternative approaches (e.g., vaccination). Furthermore, additional research is needed to clarify the biological mechanisms linking obesity and GBS colonization and to refine screening guidelines to optimize the balance between effectiveness and safety [40].

### 4.3 Strengths and Limitations

This study adheres to rigorous systematic review methodology, including comprehensive search, bias assessment, and adherence to PRISMA standards. This review’s strengths include its inclusion of diverse populations from high-, middle-, and low-income countries, as well as a broad range of study designs, including large prospective cohorts and cross-sectional studies. Many studies adjusted for relevant confounders such as age, parity, diabetes, and smoking, lending robustness to the findings. Limitations include heterogeneity in GBS testing protocols, such as differences in culture versus PCR testing, swab collection techniques and gestational timing, as well as possible residual confounding in observational data. Additionally, several studies did not adjust for socio-behavioral variables, which could confound the observed associations. Limitations in the statistical strategy used may involve the approach to diversity, which involved translating reported BMI categories into intervals on a continuous scale based on data from the Danish Medical Birth Registry. However, this allowed for a rigorous meta-analysis with results spanning a wider range of BMI values, thereby enabling the proof of dose-response associations, a strength of this review.

### 4.4 Future Directions

Further research should investigate the biological mechanisms linking adiposity to GBS colonization, particularly the roles of the microbiome, inflammation, and metabolic pathways. Additionally, explore patient perspectives and ethical implications of revised prevention protocols. Prospective cohorts integrating BMI, microbiota profiling, and inflammatory biomarkers would provide mechanistic insight. Moreover, trials are needed to assess whether lifestyle interventions, such as weight optimization or targeted probiotics, reduce GBS colonization and transmission. Lastly, modelling studies may help evaluate the cost-effectiveness and safety of incorporating BMI into GBS screening guidelines.

## 5. Conclusion

To our knowledge, this is the first systematic review and meta-analysis reporting consistent results on the association between BMI and EOGBS, suggesting an increased risk of GBS-related disease in the offspring of obese women. The integration of BMI into existing risk assessment models warrants consideration, pending further evidence and implementation analysis.

## Data Availability

All relevant data are within the manuscript and its Supporting Information files

## 6. Supporting Information

- S1 Appendix: Search Strategy
- S2 Appendix: PFO Inclusion Criteria
- S3 Appendix: PRISMA Checklist (2020)
- S4 Appendix: Leave-one-out analyses

### Support

|  |  |
| --- | --- |
| Systematic review method | Lasse Østengaard, Research Coordinator,<br>University of Southern Denmark. |
|  | Jeppe Benneskou Schroll, CEBMO (Center for Evidence-Based Medicine Odense) |
| Literature search | Peter Everfelt (Librarian, University hospital Esbjerg)<br>Lasse Østengaard, Research Coordinator, Southern University Denmark.<br>Attending course on literature search for systematic review through Southern University Denmark (Attendance EEG) |
| Quality appraisal | CEBMO (Center for Evidence-Based Medicine Odense)/Cochrane Denmark |
| Syntesis | Torben Bjerregaard Larsen, professor, MD, PhD, FESC |

### Funding

There was no financial sponsors of this work. Non-financial contributors as listed above.

---

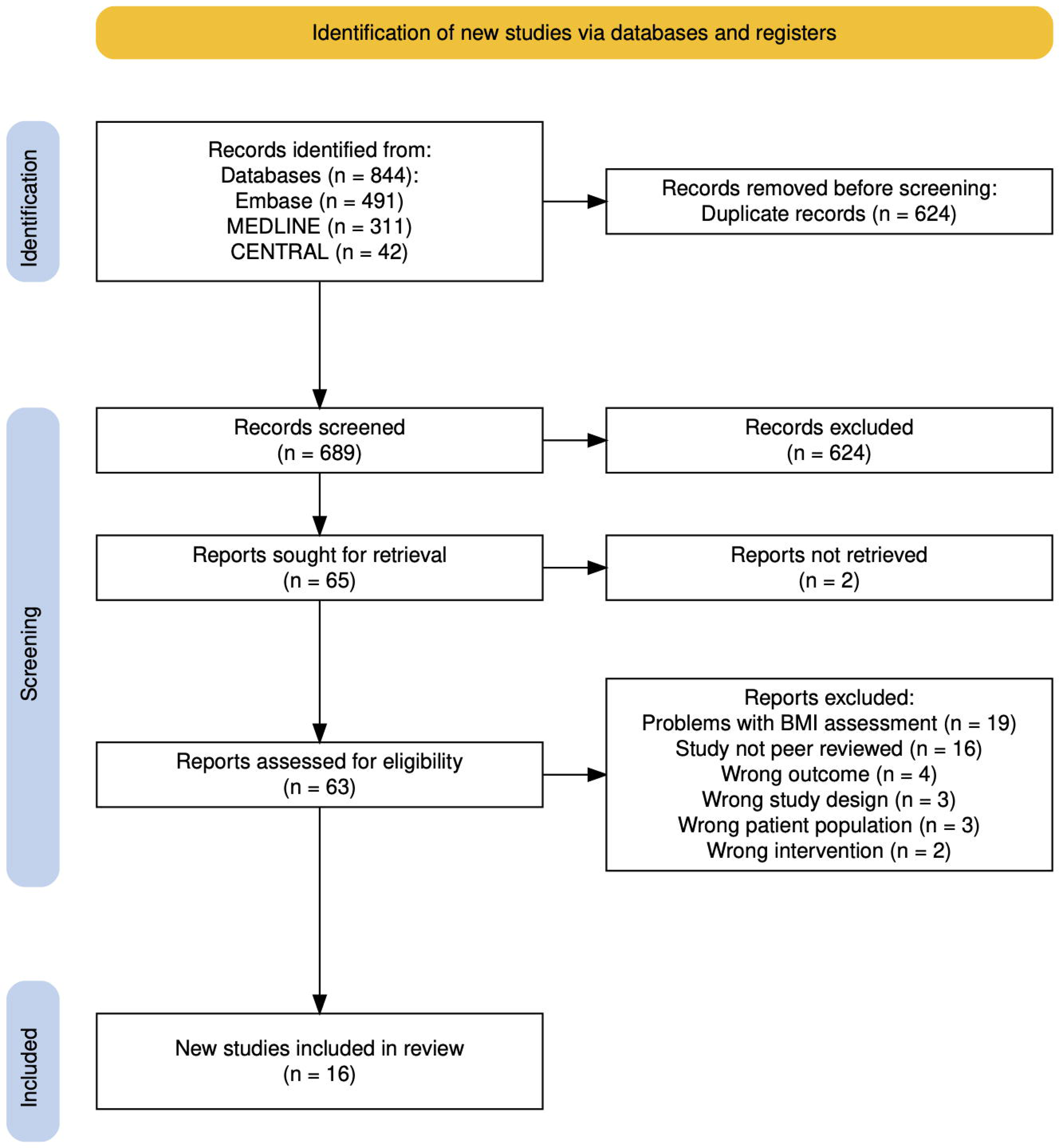

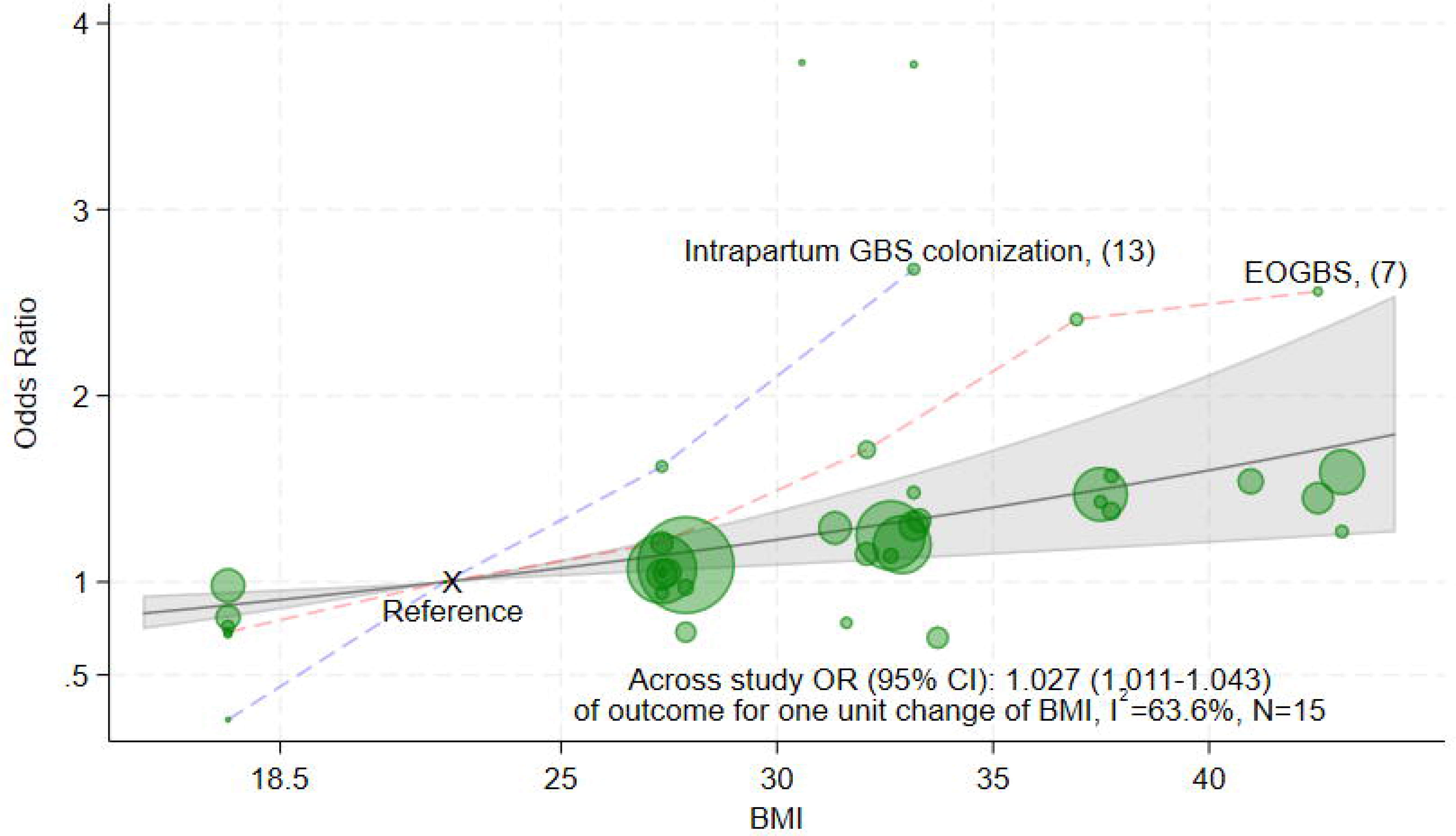

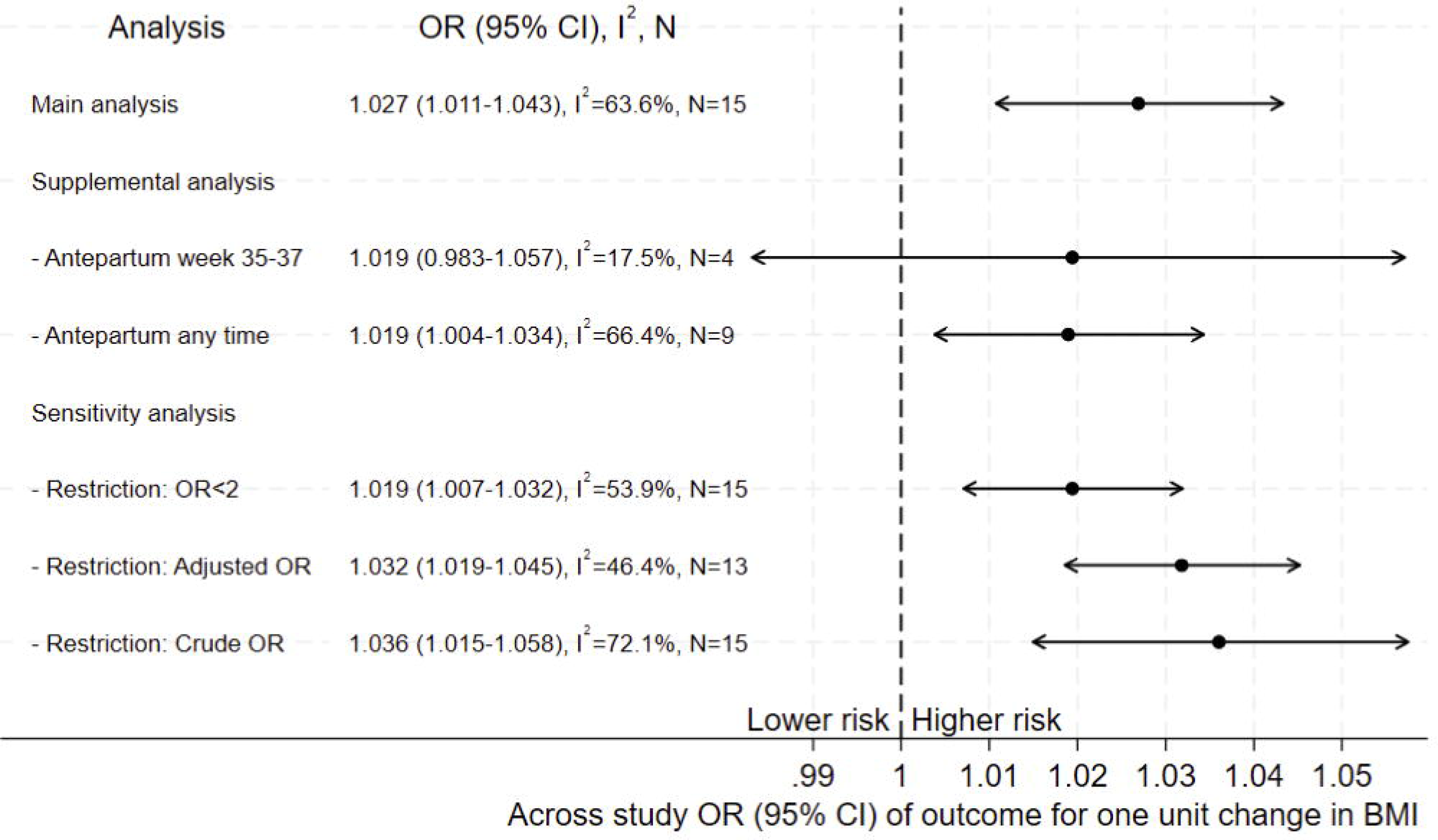

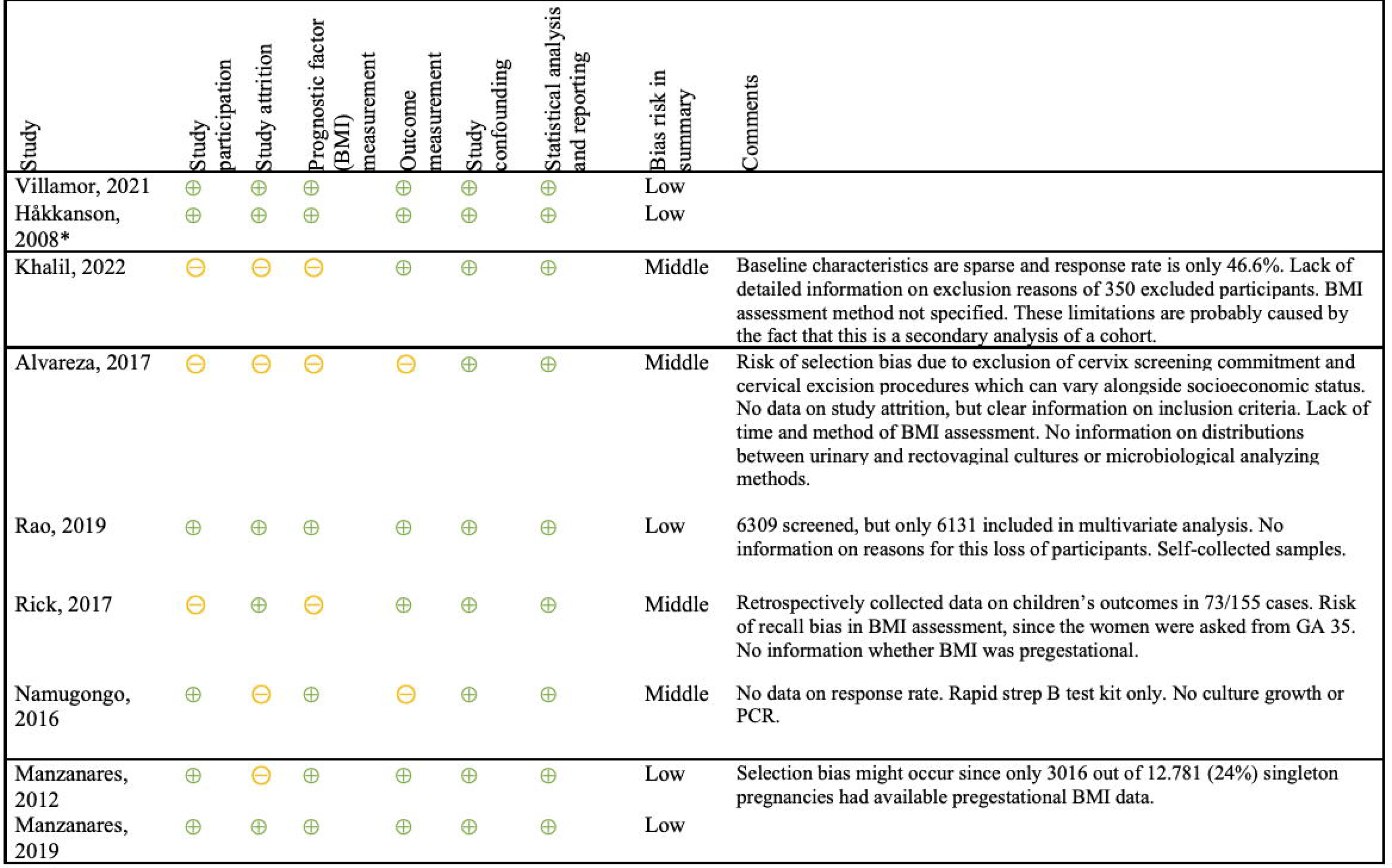

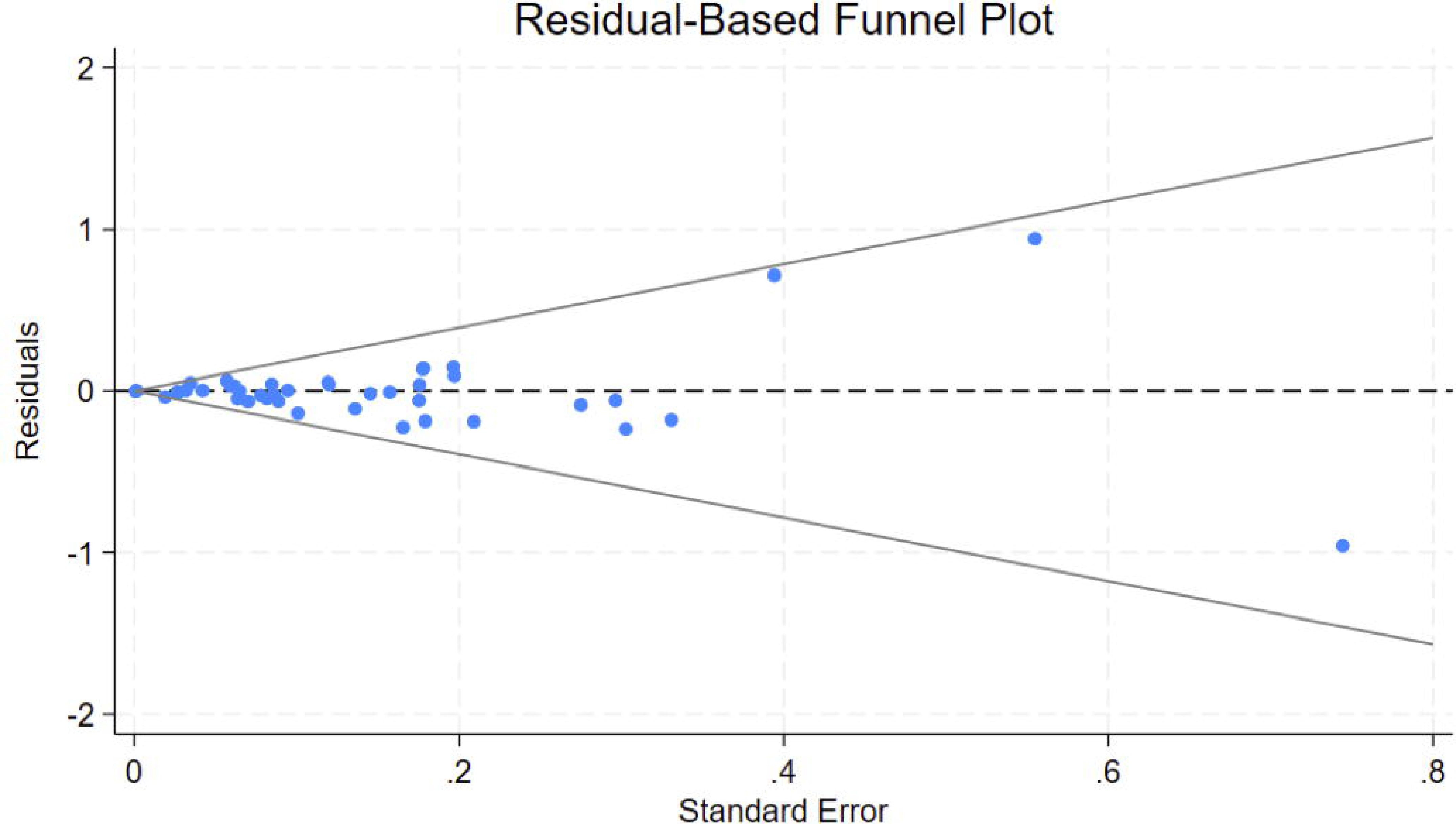

## References

1 Benitz WE, Gould JB, Druzin ML. Risk Factors for Early-onset Group B Streptococcal Sepsis: Estimation of Odds Ratios by Critical Literature Review [Internet]. 1999. Available from: http://publications.aap.org/pediatrics/article-pdf/103/6/e77/844129/e77.pdf

2 O’Sullivan CP, Lamagni T, Patel D, Efstratiou A, Cunney R, Meehan M, et al. Group B streptococcal disease in UK and Irish infants younger than 90 days, 2014–15: a prospective surveillance study. Lancet Infect Dis. 2019 Jan 1;19(1):83–90.

3 Flidel-Rimon O, Galstyan S, Juster-Reicher A, Rozin I, Shinwell ES. Limitations of the risk factor based approach in early neonatal sepsis evaluations. Acta Paediatrica, International Journal of Paediatrics. 2012 Dec;101(12).

4 Edmond KM, Scott S, Edmond KM, Kortsalioudaki C, Scott S, Schrag SJ, et al. Group B streptococcal disease in infants aged younger than 3 months: systematic review and meta-analysis. Lancet [Internet]. 2012;379:547–56. Available from: www.thelancet.com

5 Russell NJ, Seale AC, O’Driscoll M, O’Sullivan C, Bianchi-Jassir F, Gonzalez-Guarin J, et al. Maternal Colonization with Group B Streptococcus and Serotype Distribution Worldwide: Systematic Review and Meta-analyses. Vol. 65, Clinical Infectious Diseases. Oxford University Press; 2017. p. S100–11.

6 Håkansson S, Källen K. High maternal body mass index increases the risk of neonatal early onset group B streptococcal disease. Acta Paediatrica, International Journal of Paediatrics. 2008 Oct;97(10):1386–9.

7 Villamor E, Norman M, Johansson S, Cnattingius S. Maternal Obesity and Risk of Early-onset Neonatal Bacterial Sepsis: Nationwide Cohort and Sibling-controlled Studies. Clinical Infectious Diseases. 2021 Nov 1;73(9):E2656–64.

8 Venkatesh KK, Vladutiu CJ, Strauss RA, Thorp JM, Stringer JSA, Stamilio DM, et al. Association between Maternal Obesity and Group B Streptococcus Colonization in a National U.S. Cohort. J Womens Health. 2020 Dec 1;29(12):1507–12.

9 Stapleton RD, Kahn JM, Evans LE, Critchlow CW, Gardella CM. Risk Factors for Group B Streptococcal Genitourinary Tract Colonization in Pregnant Women. 2005.

10 Manzanares S, Zamorano M, Naveiro-Fuentes M, Pineda A, Rodríguez-Granger J, Puertas A. Maternal obesity and the risk of group B streptococcal colonisation in pregnant women. J Obstet Gynaecol (Lahore). 2019 Jul 4;39(5):628–32.

11 Alvareza MD, Subramaniam A, Tang Y, Edwards RK. Obesity as an independent risk factor for group B streptococcal colonization. Journal of Maternal-Fetal and Neonatal Medicine. 2017 Dec 2;30(23):2876–9.

12 Hosseini MS, Jahanshahlou F, Akbarzadeh MA, Zarei M, Vaez-Gharamaleki Y. Formulating research questions for evidence-based studies. Journal of Medicine, Surgery, and Public Health. 2024 Apr 1;2:100046.

13 Khalil MR, Hartvigsen CM, Thorsen PB, Møller JK, Uldbjerg N. Maternal age and body mass index as risk factors for rectovaginal colonization with group B streptococci. International Journal of Gynecology and Obstetrics. 2023 Apr 1;161(1):303–7.

14 Rao GG. Differential rates of group B streptococcus (GBS) colonisation in pregnant women in a racially diverse area of London, UK: a cross-sectional study. Vol. 126, BJOG: An International Journal of Obstetrics and Gynaecology. Blackwell Publishing Ltd; 2019. p. 1353.

15 Rick AM, Aguilar A, Cortes R, Gordillo R, Melgar M, Samayoa-Reyes G, et al. Group B streptococci colonization in pregnant Guatemalan women: Prevalence, Risk factors, and vaginal microbiome. Open Forum Infect Dis. 2017 Nov 1;4(1).

16 Namugongo A, Bazira J, Fajardot Y, Joseph N. Group B Streptococcus Colonization among Pregnant Women Attending Antenatal Care at Tertiary Hospital in Rural Southwestern Uganda. Int J Microbiol. 2016;2016.

17 Kleweis SM, Cahill AG, Odibo AO, Tuuli MG. Maternal obesity and rectovaginal group B streptococcus colonization at term. Infect Dis Obstet Gynecol. 2015;2015.

18 Galán SM, Hernández ÁS, Zúñiga IV, Löpez Criado MS, Lloréns AP, José Luis Gallo V. Abnormal maternal body mass index and obstetric and neonatal outcome. Journal of Maternal-Fetal and Neonatal Medicine. 2012 Mar;25(3):308–12.

19 Dahan-Saal J, Gérardin P, Robillard PY, Barau G, Bouveret A, Picot S, et al. Déterminants de la colonisation maternelle à streptocoque B et facteurs associés à sa transmission verticale périnatale: Étude cas-témoins. Gynecologie Obstetrique et Fertilite. 2011;39(5):281–8.

20 Melchor I, Burgos J, Del Campo A, Aiartzaguena A, Gutiérrez J, Melchor JC. Effect of maternal obesity on pregnancy outcomes in women delivering singleton babies: A historical cohort study. J Perinat Med. 2019 Aug 1;47(6):625–30.

21 Chen Z, Wen G, Cao X, Li S, Wang X, Yao Z, et al. Group B streptococcus colonisation and associated risk factors among pregnant women: A hospital-based study and implications for primary care. Int J Clin Pract. 2019 May 1;73(5).

22 Kwon KS, Cheng TH, Reynolds SA, Zhou J, Cai H, Lee S, et al. Maternal Group B Streptococcus Infection Correlates Inversely with Preeclampsia in Pregnant African American Women. Maternal-Fetal Medicine. 2024 Jan 1;6(1):23–8.

23 Chen Z, Wu C, Cao X, Wen G, Guo D, Yao Z, et al. Risk factors for neonatal group B streptococcus vertical transmission: a prospective cohort study of 1815 mother–baby pairs. Journal of Perinatology. 2018 Oct 1;38(10):1309–17.

24 Schuchat A, Oxtoby M, Cochi S, Sikes RK, Hightower A, Plikaytis B, et al. Population-Based Risk Factors for Neonatal Group B Streptococcal Disease: Results of a Cohort Study in Metropolitan Atlanta [Internet]. Vol. 162, Source: The Journal of Infectious Diseases. 1990. Available from: https://www.jstor.org/stable/30131700

25 Collin SM, Demirjian A, Swann C, Lamagni T. Race and Ethnicity in Neonatal Group B Streptococcal Disease in England: 2016–2020. Pediatrics. 2022 Sep 1;150(3).

26 Edwards JM, Watson N, Focht C, Wynn C, Todd CA, Walter EB, et al. Group B Streptococcus (GBS) Colonization and Disease among Pregnant Women: A Historical Cohort Study. Infect Dis Obstet Gynecol. 2019;2019.

27 Koren O, Goodrich JK, Cullender TC, Spor A, Laitinen K, Kling Bäckhed H, et al. Host remodeling of the gut microbiome and metabolic changes during pregnancy. Cell. 2012 Aug 3;150(3):470–80.

28 Eastman AJ, Moore RE, Townsend SD, Gaddy JA, Aronoff DM. The Influence of Obesity and Associated Fatty Acids on Placental Inflammation. Clin Ther. 2021 Feb 1;43(2):265–78.

29 Gaddy JA, Moore RE, Lochner JS, Rogers LM, Noble KN, Giri A, et al. Palmitate and group B Streptococcus synergistically and differentially induce IL-1β from human gestational membranes. Front Immunol. 2024;15.

30 Frieden TR, Director Harold Jaffe MW, Stephens JW, Thacker SB, Spriggs Terraye M Starr SR, Doan QM, et al. Morbidity and Mortality Weekly Report Prevention of Perinatal Group B Streptococcal Disease [Internet]. 2010. Available from: www.cdc.gov/mmwrhttp://www.cdc.gov/mmwr/cme/conted.html

31 Denison FC, Aedla NR, Keag O, Hor K, Reynolds RM, Milne A, et al. Care of Women with Obesity in Pregnancy: Green-top Guideline No. 72. BJOG. 2019 Feb 1;126(3):e62–106.

32 Neonatal infection: antibiotics for prevention and treatment NICE guideline [Internet]. 2021. Available from: www.nice.org.uk/guidance/ng195

33 Mercado-Evans V, Zulk JJ, Hameed ZA, Patras KA. Gestational diabetes as a risk factor for GBS maternal rectovaginal colonization: a systematic review and meta-analysis. BMC Pregnancy Childbirth. 2024 Dec 1;24(1).

34 Rosenberg R, Normann L;, Katrine A, Henriksen ;, Fenger-Grøn B;, Møller J;, et al. Risk-based screening and intrapartum group B streptococcus polymerase chain reactionresults reduce use of antibiotics during labour [Internet]. Available from: https://ugeskriftet.dk/dmj/risk-

35 Hartvigsen CM, Nielsen SY, Møller JK, Khalil MR. Reduction of intrapartum antibiotic prophylaxis by combining risk factor assessment with a rapid bedside intrapartum polymerase chain reaction testing for group B streptococci. European Journal of Obstetrics & Gynecology and Reproductive Biology. 2022 May 1;272:173–6.

36 Ramesh Babu S, McDermott R, Farooq I, Le Blanc D, Ferguson W, McCallion N, et al. Screening for group B Streptococcus (GBS) at labour onset using PCR: accuracy and potential impact–a pilot study. J Obstet Gynaecol (Lahore). 2018 Jan 2;38(1):49–54.

37 https://www.esundhed.dk/Emner/Graviditet-foedsler-og-boern/Nyfoedte-og-foedsler-1997.

38 Towers C V., Rumney PJ, Asrat T, Preslicka C, Ghamsary MG, Nageotte MP. The accuracy of late third-trimester antenatal screening for group B streptococcus in predicting colonization at delivery. Am J Perinatol. 2010;27(10):785–9.

39 Hussain FN, Al-Ibraheemi Z, Pan S, Francis AP, Taylor D, Lam MC, et al. The Accuracy of Group Beta Streptococcus Rectovaginal Cultures at 35 to 37 Weeks of Gestation in Predicting Colonization Intrapartum. AJP Rep. 2019;9(3):E302–9.

40 Giorgakoudi K, O’Sullivan C, Heath PT, Ladhani S, Lamagni T, Ramsay M, et al. Cost-effectiveness analysis of maternal immunisation against group B Streptococcus (GBS) disease: A modelling study. Vaccine. 2018 Nov 12;36(46):7033–42.

