## Supplementary material for "Maternal Body Mass Index and the Risk of Early-Onset Group B Streptococcus Disease in Newborns: A Systematic Review and Meta-Analysis": S1 Appendix (resubmission). Search strategy .pdf

Search string from Medline, CENTRAL and Embase Classic+Embase <1947 to 2024 August 5>

| | EMBASE | Medline | CENTRAL |
|---|---|---|---|
| 1 | exp high risk pregnancy/ | exp Pregnancy, HighRisk/ | MeSH descriptor: [Pregnancy] explode all trees |
| 2 | exp pregnancy/ | exp Pregnancy/ | MeSH descriptor: [Pregnancy Complications] explode all trees |
| 3 | exp Pregnancy Complications/ | exp Pregnancy Complications/ | MeSH descriptor: [Pregnancy, High-Risk] explode all trees |
| 4 | exp childbirth/ | exp Parturition/ | MeSH descriptor: [Parturition] explode all trees |
| 5 | "maternal".ti,ab,kf. | "pregnan*".ti,ab,kf. | ("pregnancy"):ti,ab,kw (Word variations have been searched) |
| 6 | pregnan*.ti,ab,kf. | "maternal".ti,ab,kf. | (maternal):ti,ab,kw (Word variations have been searched) |
| 7 | (childbirth or partus or birth or lab?r or obstetric).ti,ab,kf. | (childbirth or partus or birth or lab?r or obstetric).ti,ab,kf. | (childbirth or partus or birth or labor or labour or obstetric):ti,ab,kw |
| 8 | 1 OR 2 OR 3 OR 4 OR 5 OR 6 OR 7 | 1 OR 2 OR 3 OR 4 OR 5 OR 6 OR 7 | 1 OR 2 OR 3 OR 4 OR 5 OR 6 OR 7 |
| 9 | exp morbid obesity/ | exp Body Weight/ | MeSH descriptor: [Obesity] explode all trees |
| 10 | exp Body Mass Index/ | exp Obesity, Morbid/ | MeSH descriptor: [Pregnancy in Obesity] explode all trees |
| 11 | exp maternal obesity/ | exp Obesity, Maternal/ | MeSH descriptor: [Obesity, Morbid] explode all trees |
| 12 | exp obesity/ | exp Obesity/ | MeSH descriptor: [Body Mass Index] explode all trees |
| 13 | exp body mass/ | exp Body Mass Index/ | MeSH descriptor: [Ideal Body Weight] explode all trees |
| 14 | "obes*".ti,ab,kf. | "BMI".ti,ab,kf. | (overweight):ti,ab,kw (Word variations have been searched) |
| 15 | "BMI".ti,ab,kf. | "obes*".ti,ab,kf. | (obesity):ti,ab,kw (Word variations have been searched) |
| 16 | "overweight".ti,ab,kf. | "overweight".ti,ab,kf. | (BMI):ti,ab,kw (Word variations have been searched) |
| 17 | (adipositas or adiposity or body weight excess or corpulency or obesitas or fat mass or body weight).ti,ab,kf. | (body mass index or body mass or maternal obesity or morbid obesity).ti,ab,kf. | (adipositas or adiposity or body weight excess or corpulency or obesitas or fat mass or body weight):ti,ab,kw (Word variations have been searched) |
| 18 | (body mass index or body mass or maternal obesity or morbid obesity).ti,ab,kf. | (adipositas or adiposity or body weight excess or corpulency or obesitas or fat mass or body weight).ti,ab,kf. | (body mass index or body mass or maternal obesity or morbid obesity):ti,ab,kw (Word variations have been searched) |
| 19 | 9 OR 10 OR 11 OR 12 OR 13 OR 14 OR 15 OR 16 OR 17 OR 18 | 9 OR 10 OR 11 OR 12 OR 13 OR 14 OR 15 OR 16 OR 17 OR 18 | 9 OR 10 OR 11 OR 12 OR 13 OR 14 OR 15 OR 16 OR 17 OR 18 |
| 20 | exp Streptococcal infections/ | exp Streptococcal Infections/ | MeSH descriptor: [Streptococcus agalactiae] explode all trees |
| 21 | exp Streptococcus agalactiae/ | exp Streptococcus agalactiae/ | MeSH descriptor: [Streptococcal Infections] explode all trees |

| | | | |
|---|---|---|---|
| 22 | (group b streptococc* or beta h* streptococcus group b or ha?molytic streptococcus b or hemolytic streptococcus b or staphylococc* agalactiae or streptococcus group B or streptococcus group b or streptococc* | (group b streptococc* or beta h* streptococcus group b or ha?molytic streptococcus b or hemolytic streptococcus b or staphylococc* agalactiae or | (group b streptococcus OR beta h streptococcus group b OR haemolytic streptococcus b OR staphylococcus agalactiae):ti,ab,kw (Word variations have been searched) |
| | agalactia* or streptococc* infection* or streptococc* coloni?ation* or GBS coloni?ation* or GBS infection* or (GBS adj3 screening) or (streptococc* adj3 b) or streptococc*).ti,ab,kf. | streptococcus group B or streptococcus group b or streptococc* agalactia* or streptococc* infection* or streptococc* coloni?ation* or GBS coloni?ation* or GBS infection* or (GBS adj3 screening) or (streptococc* adj3 b) or streptococc*).ti,ab,kf. | |
| 23 | ("Early Onset GBS" or EOGBS).ti,ab,kf. | ("Early Onset GBS" or EOGBS).ti,ab,kf. | (Early Onset GBS OR EOGBS):ti,ab,kw (Word variations have been searched) |
| 24 | 20 OR 21 OR 22 OR 23 | 20 OR 21 OR 22 OR 23 | (group b streptococc* or beta h* streptococcus group b or haemolytic streptococcus b or hemolytic streptococcus b or staphylococc* agalactiae or streptococcus group B or streptococcus group b or streptococc* agalactia* or streptococc* infection* or streptococc* colonisation* or GBS colonisation* or GBS infection* or (GBS adj3 screening) or (streptococc* adj3 b) or streptococc*):ti,ab,kw (Word variations have been searched) |
| 25 | 8 AND 19 AND 24 | 8 AND 19 AND 24 | 20 OR 21 OR 22 OR 23 OR 24 |
| | | | 8 AND 19 AND 25 |
