## Supplementary material for "Maternal Body Mass Index and the Risk of Early-Onset Group B Streptococcus Disease in Newborns: A Systematic Review and Meta-Analysis": S2 Appendix. PFO Inclusion Criteria.docx

| **POPULATION** | Pregnant women |
| --- | --- |
| **PROGNOSTIC FACTOR** | Primary: Pre-gestational (in this study acceptance of BMI assessment before 10^th^ gestational week) overweight/obese (BMI ≥ 25)  Secondary: BMI categories:  25–29.9  30-34,9  35-39,9  ≥ 40 |
| **COMPARATOR(S)/CONTROL** | Normal weight (BMI 18,5-24,9) |
| **MAIN OUTCOME(S)**  **MEASURES OF EFFECT** | EOGBS and its proxy outcomes, including; intrapartum vGBS, and rectovaginal or urinary GBS colonization before term  Based on outcome data (e.g. odds ratios (OR) and risk ratios (RR)) |
| **TYPES OF STUDY TO BE INCLUDED** | Cohort studies, cross-sectional studies, RCT, case control studies |
