## Supplementary figures and images for "Maternal Body Mass Index and the Risk of Early-Onset Group B Streptococcus Disease in Newborns: A Systematic Review and Meta-Analysis"

### S4 Appendix. Leave-one-out analyses.pdf

**S6 Appendix** Leave out analysis

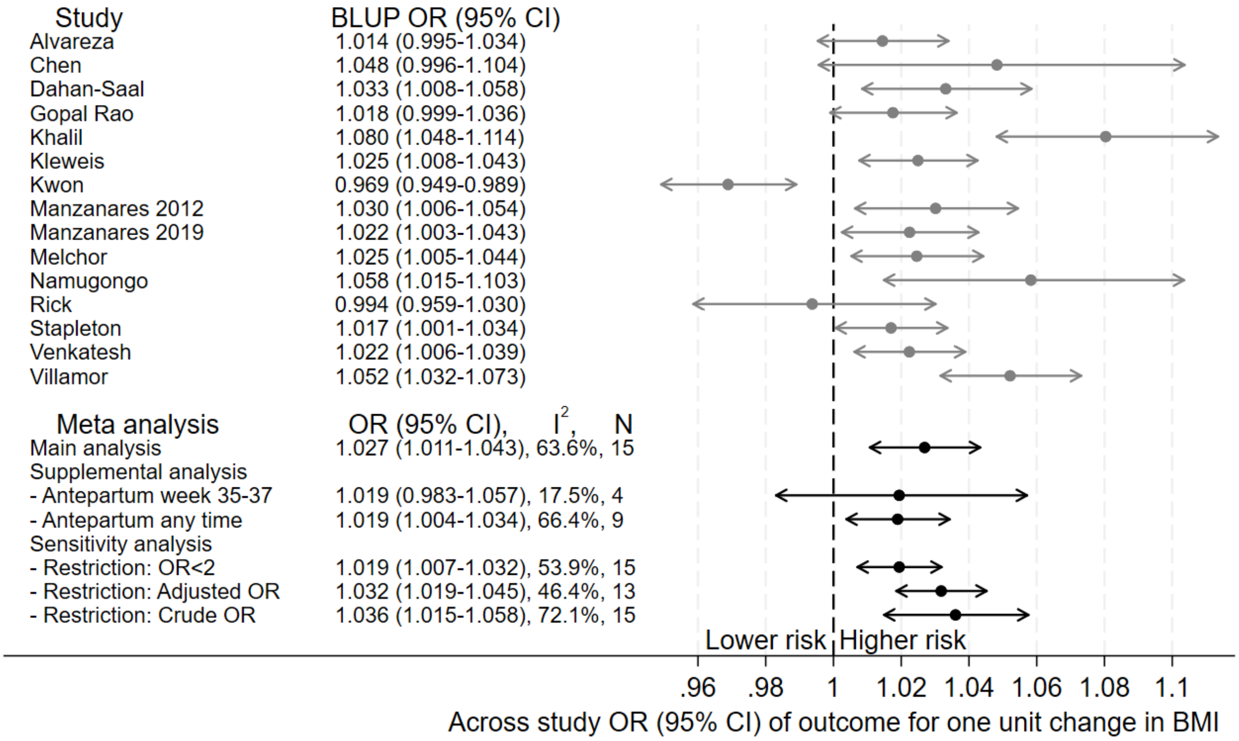
